# Scaling-up Cervical Cancer Services for Women Living with HIV in Mozambique, October 2018–September 2023

**DOI:** 10.1101/2024.11.21.24317677

**Authors:** Della Correia, Zurnaid Bay, Argentina Wate, Herminio Nhanguiombe, Pinto Manhica, Erica Bila, Sheila Tualufo, Celina Mate, Celeste Amado, Cesaltina Lorenzoni

**Author notes:** **Corresponding Author:** Zurnaid Bay.

## Abstract

**Background:** Cervical cancer is the most common cancer among women in Mozambique. Women living with HIV (WLHIV) have a six times greater risk of developing cervical cancer. National guidelines call for annual screening of all HIV-positive women aged 20-49 years or below 20 years if she is sexually active and HIV-negative women aged 25-49 years, with follow-up after 1 year for HIV-positive and 3 years for HIV-negative women that screen negative. Women screened positive are referred to treatment with cryotherapy, thermal ablation, or loop electrosurgical excision procedure (LEEP). In 2021, HIV prevalence among women aged 15–49 years was estimated at 15.4%. We analyzed routine program data to describe the expansion of cervical cancer services and identify gaps in screening and treatment coverage among WLHIV since 2018.

**Methods:** We analyzed semi-annual, routinely collected programmatic data on cervical cancer screening and treatment services reported between 21 September 2018 to 20 March 2019 (585 sites reported) and 21 March to 20 September 2023 (610 sites reported). We verify rates of screening coverage (WLHIV screened divided by WLHIV on ART aged ≥15 years), screen positivity (WLHIV screened positive divided by number screened), and treatment (WLHIV treated for precancerous lesions divided by number screened positive).

**Results:** The screening coverage improved from 1.4% (10,596 screened/731,572 WLHIV on ART) in October 2018–March 2019 to 36.2% (395,609/1,094,033) in March–September 2023. In the same period, screen positivity rates increased from 8.1% (861/10,596) to 10.9% (43,290/395,614), and treatment rates increased from 59.9% (516/861) to 94.0% (40,677/43,290).

**Conclusion:** Access to cervical cancer services has improved dramatically for a large population of WLHIV. With increased screening, more WLHIV with pre-cancerous lesions have been found and treated, averting morbidity and mortality due to cervical cancer. Quality improvement strategies may further improve service delivery as the program expands to reach all eligible WLHIV in Mozambique.

## Introduction

Globally, though preventable and curable if detected early, cervical cancer is the fourth most common cancer among women, with almost 660,000 new cases and 350,000 deaths in 2022.^1^ Nearly 94% of cervical cancer deaths occurred in low- and middle-income countries.^1^ Among women living with HIV (WLHIV), cervical cancer is the most common cancer, and WLHIV are six times more likely to develop invasive cervical cancer compared to women without HIV infection.^1^

Mozambique has a population of 16.5 million women aged ≥15 years who are at risk of developing cervical cancer. At 33.4%, Mozambique has one of the highest incidence rates of cervical cancer in the world, and cervical cancer is the leading cause of cancer-related mortality among women aged ≥15 years in Mozambique.^2,3^

The Mozambique national cervical and breast cancer program was established in 2009 to provide prevention and early treatment of precancerous lesions among women and to reduce the incidence of and mortality due to those cancers.^4^ Under the national guidelines, the target group for cervical cancer screening and treatment is HIV-negative women aged 25–49 years and all HIV-positive women aged 20-49 years or below 20 years if she is sexually active. Since 2009, the Ministry of Health of Mozambique (MISAU) has recommended visual inspection with acetic acid (VIA) for screening.^4^ Additionally, where testing capacity is available, specimens may be tested for human papillomavirus (HPV) using GeneXpert. Screening is conducted using VIA or HPV testing. VIA screening results can be positive, negative, or suspected cancer. “Screen-positive” indicates the presence of an aceto-white lesion on the cervix or a positive HPV test using GeneXpert, and only those women who test HPV positive should receive VIA. Therefore, all women with aceto-white lesions smaller than 75% of the cervix (cervical mucosa that changed color to white after acetic acid application), as well as WLHIV who were negative by VIA but tested positive for HPV serotype 16 or 18, were eligible for treatment with cryotherapy or thermal ablation. WLHIV, who tested positive for other HPV serotypes and VIA negative, are recommended for follow-up in one year. MISAU recommends treatment with LEEP for acetowhite lesions larger than 75% of the cervix identified by VIA screening and referral to a reference center for diagnosis, management, and treatment of suspected invasive cancers.

MISAU recommends rescreening annually for WLHIV and every 3 years for HIV-negative women.^4^ Although HIV-negative women are outside the scope of this paper, they are also at risk for cervical cancer, and because of this risk, they should be screened every 3 years according to Mozambique guidelines.^4^ In alignment with global objectives, MISAU created a national roadmap toward cervical cancer elimination as a public health threat by 2030 to guide Mozambique in reaching the 2030 global 90-70-90 targets (90% of eligible girls vaccinated against HPV; 70% of women screened for precancerous lesions of the cervix with a high-performance test; and 90% of precancerous lesions treated/90% of women with invasive cancer properly managed).^5^

The objective of this manuscript is to describe the expansion of cervical cancer services and identify gaps in coverage and treatment for WLHIV from 2018 and 2023.

## Methods

We conducted a retrospective analysis of routinely reported Monitoring, Evaluation, and Reporting (MER) data from sites supported by the U.S. President’s Emergency Plan for AIDS Relief (PEPFAR) reporting cervical cancer screening and treatment services among WLHIV between 21 September 2018 to 20 March 2019 (585 sites reported) and 21 March to 20 September 2023 (610 sites reported). The MER data were collected and aggregated at the facility level and entered into the Data for Accountability, Transparency, and Impact (DATIM) system semi-annually by PEPFAR implementing partners (IPs). Descriptive analysis was performed using Excel.

WLHIV on ART is defined as women ≥15 years old living with HIV on ART. We defined screening coverage as the number of WLHIV screened divided by the total number of WLHIV on ART aged ≥15 years, and the screen positivity rate as the number of WLHIV who had a positive screen by VIA or HPV testing over the total number of WLHIV who were screened.

Treatment was defined as any treatment (e.g., cryotherapy, thermal ablation, or LEEP) for precancer cervical lesions following screening positive. The treatment rate was defined as the proportion of WLHIV who received treatment among those who screened positive.^1^

We analyzed data by time period, province, and treatment type (e.g., cryotherapy, thermal ablation, LEEP).

## Results

In September 2018, there were 731,572 WLHIV aged ≥15 years on ART in Mozambique. From October 2018 to September 2023, 1,460,904 screenings were performed among WLHIV aged ≥15 years. Screening coverage increased from 1.4% (10,596 screened/731,572 WLHIV on ART) in October 2018–March 2019 to 36,2 % (395.609/1.094.033) in March–September 2023 (Table 1). These periods represent the first 6 month reporting period compared to the last 6 months.

**Table 1:**
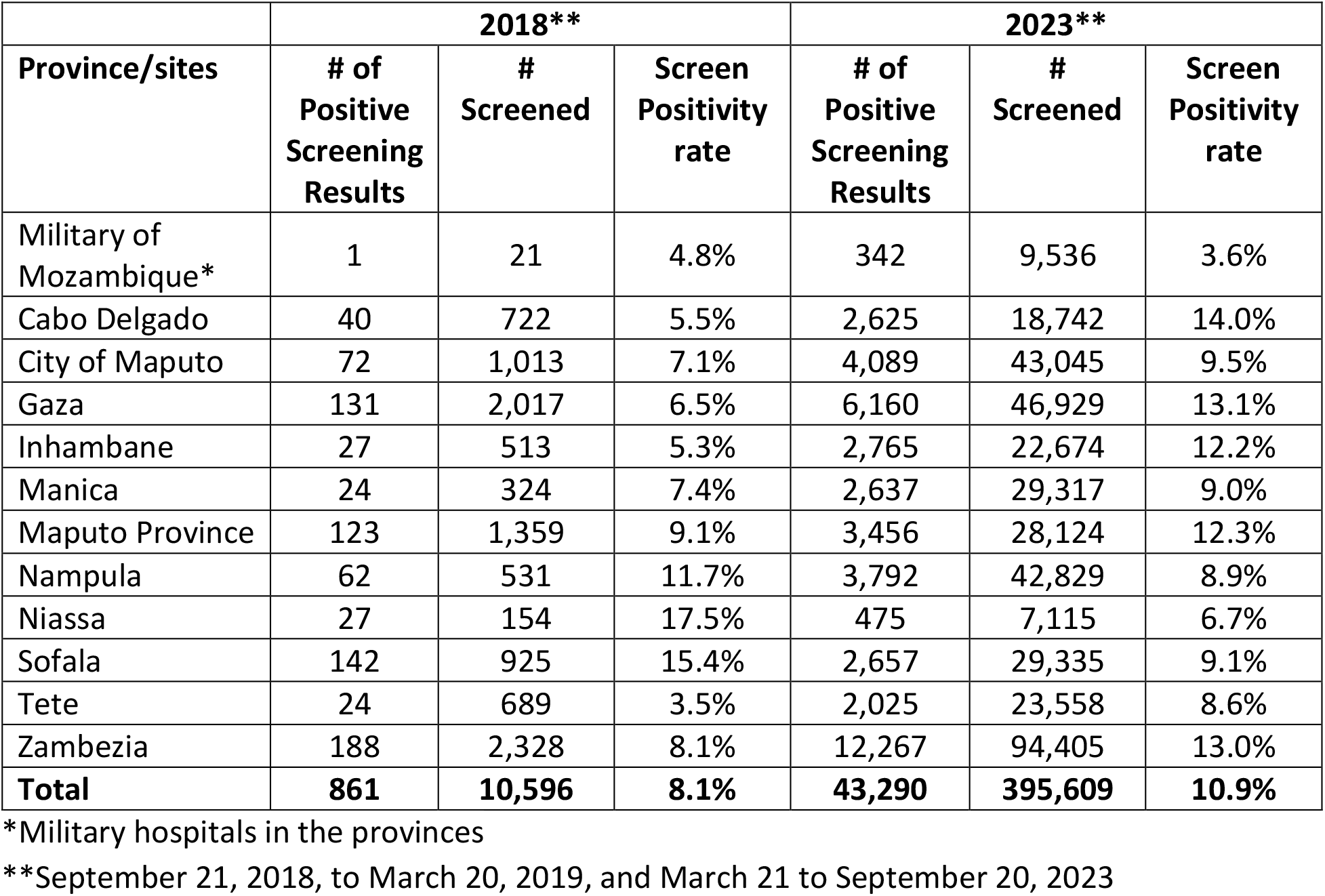
Precancer Screening Positivity Rates by Province among WLHIV at PEPFAR-Supported Sites in Mozambique, 2018** e 2023**.

| Province/sites | 2018** |  |  | 2023** |  |  |
| --- | --- | --- | --- | --- | --- | --- |
|  | # of Positive Screening Results | # Screened | Screen Positivity rate | # of Positive Screening Results | # Screened | Screen Positivity rate |
| Military of Mozambique* | 1 | 21 | 4.8% | 342 | 9,536 | 3.6% |
| Cabo Delgado | 40 | 722 | 5.5% | 2,625 | 18,742 | 14.0% |
| City of Maputo | 72 | 1,013 | 7.1% | 4,089 | 43,045 | 9.5% |
| Gaza | 131 | 2,017 | 6.5% | 6,160 | 46,929 | 13.1% |
| Inhambane | 27 | 513 | 5.3% | 2,765 | 22,674 | 12.2% |
| Manica | 24 | 324 | 7.4% | 2,637 | 29,317 | 9.0% |
| Maputo Province | 123 | 1,359 | 9.1% | 3,456 | 28,124 | 12.3% |
| Nampula | 62 | 531 | 11.7% | 3,792 | 42,829 | 8.9% |
| Niassa | 27 | 154 | 17.5% | 475 | 7,115 | 6.7% |
| Sofala | 142 | 925 | 15.4% | 2,657 | 29,335 | 9.1% |
| Tete | 24 | 689 | 3.5% | 2,025 | 23,558 | 8.6% |
| Zambezia | 188 | 2,328 | 8.1% | 12,267 | 94,405 | 13.0% |
| **Total** | **861** | **10,596** | **8.1%** | **43,290** | **395,609** | **10.9%** |

\*Military hospitals in the provinces
\*\*September 21, 2018, to March 20, 2019, and March 21 to September 20, 2023

In the same period, screen positivity rates increased from 8.1% (861/10,596; range = 3.5% [Tete Province] to 17.5% [Niassa Province]) to 10.9% (43,290/395,614; range = 3.6% [Military-supported sites] to 14.0% [Cabo Delgado Province]) (Table 1); treatment rates increased from 59.9% (516/861; range = 0.0% [Tete Province] to 100% [Military-supported sites]) to 94.0% (40,677/43,290; range = 89.7% [Maputo Province] to 99.2% [Niassa Province]) (Table 2). The treatment rate for cervical cancer in the last 5 years has almost doubled.

**Table 2:** Treatment Rate by Province among WLHIV at PEPFAR-Supported Sites in Mozambique, 2018** and 2023**.

|  | **2018\*\*** |  |  | **2023\*\*** |  |  |
| --- | --- | --- | --- | --- | --- | --- |
| **Province/sites** | **# of treated** | **# of Positive Screening** | **Treatment rate** | **# of treated** | **# of Positive Screening** | **Treatment rate** |
| Military of Mozambique* | 1 | 1 | 100.0% | 327 | 342 | 95.6% |
| Cabo Delgado | 11 | 40 | 27.5% | 2,479 | 2,625 | 94.4% |
| City of Maputo | 53 | 72 | 73.6% | 3,762 | 4,089 | 92.0% |
| Gaza | 126 | 131 | 96.2% | 5,940 | 6,160 | 96.4% |
| Inhambane | 17 | 27 | 63.0% | 2,623 | 2,765 | 94.9% |
| Manica | 1 | 24 | 4.2% | 2,603 | 2,637 | 98.7% |
| Maputo Province | 67 | 123 | 54.5% | 3,099 | 3,456 | 89.7% |
| Nampula | 59 | 62 | 95.2% | 3,596 | 3,792 | 94.8% |
| Niassa | 24 | 27 | 88.9% | 471 | 475 | 99.2% |
| Sofala | 29 | 142 | 20.4% | 2,455 | 2,657 | 92.4% |
| Tete | 0 | 24 | 0.0% | 1,952 | 2,025 | 96.4% |
| Zambezia | 128 | 188 | 68.1% | 11,370 | 12,267 | 92.7% |
| **Total** | **516** | **861** | **59.9%** | **40,677** | **43,290** | **94.0%** |

\*Military hospitals in the provinces
\*\*September 21, 2018, to March 20, 2019, and March 21 to September 20, 2023

In 2018, cryotherapy was used for 96.1% of all cervical precancer treatment conducted among eligible women at PEPFAR-supported sites reporting (496/516), and the remaining 3.9% of treatment utilized LEEP (20/516); the province with the greatest proportion of WLHIV treated by LEEP was Nampula Province (18.6% [11/59]). In 2023, thermal ablation made up 63.1% of all cervical cancer treatment among the same population (25,673/40,677), cryotherapy was 33.3% (13,541/40,677), and LEEP was 3.6% (1,463/40,677); the province with the greatest proportion of WLHIV treated by LEEP was Cabo Delgado Province (6.9% [170/2,479]) (Table 3).

**Table 3:** Cervical Cancer Treatment Method among WLHIV at PEPFAR-Supported Sites in Mozambique, 2018** and 2023**.

|  | **2018\*\*** |  |  | **2023\*\*** |  |  |
| --- | --- | --- | --- | --- | --- | --- |
| **Province/sites** | **Cryotherapy** | **LEEP** | **Thermal Ablation** | **Cryotherapy** | **LEEP** | **Thermal Ablation** |
| Military Mozambique* | 1 | 0 | 0 | 187 | 2 | 138 |
| Cabo Delgado | 11 | 0 | 0 | 874 | 170 | 1,435 |
| City of Maputo | 53 | 0 | 0 | 279 | 148 | 3,335 |
| Gaza | 126 | 0 | 0 | 2,427 | 312 | 3,201 |
| Inhambane | 17 | 0 | 0 | 1,278 | 62 | 1,283 |
| Manica | 1 | 0 | 0 | 1,630 | 80 | 893 |
| Maputo Province | 67 | 0 | 0 | 1,058 | 168 | 1,873 |
| Nampula | 48 | 11 | 0 | 1,305 | 73 | 2,218 |
| Niassa | 24 | 0 | 0 | 221 | 21 | 229 |
| Sofala | 29 | 0 | 0 | 1,158 | 88 | 1,209 |
| Tete | 0 | 0 | 0 | 1,113 | 19 | 820 |
| Zambezia | 119 | 9 | 0 | 2,011 | 320 | 9,039 |
| **Total** | **496** | **20** | **0** | **13,541** | **1,463** | **25,673** |

\*Military hospitals in the provinces
\*\*September 21, 2018, to March 20, 2019, and March 21 to September 20, 2023

## Discussion

Access to cervical cancer screening and treatment services among WLHIV has dramatically expanded in Mozambique from 2018 to 2023, with screening coverage reaching almost one-third of age-eligible WLHIV and treatment rates reaching nearly 100%. Improvements in access are largely due to cervical cancer screening integration into HIV services, health providers’ training, and expansion of thermal ablation treatment. Increased cervical cancer screening and treatment for precancerous lesions can avert morbidity and mortality due to cervical cancer among women with HIV.

Despite progress, gaps in cervical cancer screening remain, like in other countries.^6^ Although all WLHIV are eligible for annual screening, less than 50% of WLHIV were screened by 2023 in PEPFAR sites. On the other hand, coverage estimates may be low because of the inclusion of all 15–19-year-olds in the denominator, even though they do not need screening if they are not sexually active. Quality of screening is also a concern as the program has rapidly grown, and although health professionals have been trained, continuous mentoring and supervision are crucial. The screening using VIA is subjective which can lead to both under- and over-screening positivity rates.^7^ In Mozambique, HPV DNA testing is not widely available due to cost and infrastructure, although WHO recommends HPV DNA testing over VIA where possible.^6,8^ Mozambique’s cervical cancer program launched the roadmap for the elimination of cervical cancer as a public health priority by 2030, which will be an opportunity to address these gaps described above.^5^

The number of eligible WLHIV who had access to cervical cancer treatment over the past 5 years has increased 80-fold and surpassed the 90% WHO recommended target against the total WLHIV diagnosed with precancerous lesions. The rapid introduction of thermal ablation in 2021-2022 following the WHO recommendation has played an important role in the rapid expansion of treatment. Thermal ablation is as safe and effective as cryotherapy. The Mozambique team prioritized allocation of the thermal ablation equipment in high-volume sites to reach a higher volume of women, taking advantage of the technology being a lightweight, portable, battery-operated device requiring short treatment duration when compared to cryotherapy, which requires a supply of refrigerant gas and has a relatively long treatment duration. Therefore, the rapid expansion of thermal ablation benefited the Mozambican program. The same has yet to happen with the LEEP treatment reserved for women with lesions above 75%. Mozambique is a large country, and women referred for treatment often travel long distances to reach the few LEEP treatment centers.

One limitation of this report is that the data source only includes sites supported by PEPFAR (610 out of 1807 total health facilities). Additionally, during the analysis period (2018-2023), older MISAU cervical cancers registers were in use, and each implementing partner used a non-standardized process for entering data into a spreadsheet and transcribing it into DATIM, which may have negatively impacted data quality during this period. Lastly, the available data were aggregated at the provincial level, and additional statistical analyses were not possible.

## Conclusion

Access to cervical cancer services has improved dramatically for a large population of WLHIV. With increased screening, more WLHIV with pre-cancerous lesions have been found and treated, averting morbidity and mortality due to cervical cancer. Quality improvement strategies may improve service delivery as the program further expands to reach all eligible WLHIV in Mozambique.

## Data Availability

All data produced are available from PEPFAR Monitoring, Evaluation, and Reporting program data.

https://data.pepfar.gov/datasets#PDD

## Acknowledgments

We thank Maggie Gutierrez-Nkomo for her assistance and support. We also thank Maria Ines de Deus and Emilio Dirlikov for their ideas and encouragement.

## Funding Acknowledgement

This manuscript has been supported by the President’s Emergency Plan for AIDS Relief (PEPFAR) through the Centers for Disease Control and Prevention (CDC). The findings and conclusions in this manuscript are those of the author(s) and do not necessarily represent the official position of the funding agencies.

## Footnotes

1 This activity was reviewed by CDC, deemed not research, and was conducted consistent with applicable federal law and CDC policy. See e.g., 45 C.F.R. part 46.102(l)(2), 21 C.F.R. part 56; 42 U.S.C. §241(d); 5 U.S.C. §552a; 44 U.S.C. §3501 et seq.

